# Effects of Race on Chronic Pain in a Randomized Clinical Trial of Integrative Medical Group Visits

**DOI:** 10.1101/2023.03.03.23286767

**Authors:** Justin J. Polcari, Angela C. Incollingo Rodriguez, Benjamin C. Nephew, Veronica Melican, Jean A. King, Paula Gardiner

## Abstract

Chronic pain is one of the most common reasons adults seek medical care in the US, with estimates of prevalence ranging from 11% to 40% and relatively higher rates in diverse populations. Mindfulness meditation has been associated with significant improvements in pain, depression, physical and mental health, sleep, and overall quality of life. Group medical visits are increasingly common and are effective at treating myriad illnesses including chronic pain. Integrative Medical Group Visits (IMGV) combine mindfulness techniques, evidence based integrative medicine, and medical group visits and can be used as adjuncts to medications, particularly in diverse underserved populations with limited access to non-pharmacological therapies. The objective of the present study was to assess the effects of race on the primary pain outcomes and evaluate potential relationships between race and additional patient characteristics in data from a randomized clinical trial of IMGV in socially diverse, marginalized patients suffering from chronic pain and depression. It was hypothesized that there would be racial differences in the effects of IMGV on pain outcomes. Our analyses identified significant racial differences in the response to IMGV. Black subjects had increased pain severity throughout the duration of the 21-week study but were less likely to respond to the pain intervention compared to White subjects. These results may be related to differential comorbidity rates, catastrophizing, and digital health literacy among these participant groups. To improve patient outcomes in similar studies, interactions between pain outcomes and these factors require further investigation to affect levels and trajectory of pain severity and enhance the response to complimentary interventions.

## INTRODUCTION

Chronic pain is one of the most common reasons adults seek medical care in the United States (U.S.), with estimates of chronic pain prevalence ranging from 11% to 40% and relatively higher rates in historically under represented populations (Dahlhamer, Lucas et al. 2018). Patients with chronic pain are more likely to use health care services than the general population, which increases associated medical costs. Chronic pain is also associated with overall poor health and several significant comorbidities, such as depression and insomnia, (Morales and Yong 2021, Overstreet, Pester et al. 2023).

Previous research indicates that residency in racially minoritized and low-income neighborhood is associated with higher rates of chronic disease including chronic pain (Webster, Rice et al. 2019). Accordingly, addressing social determinants of health has been proffered as a critical step in improving overall health, especially with regards to chronic pain (Webster, Rice et al. 2019). Additionally, findings from a 12-year longitudinal study highlight the importance of considering socioeconomic status when assessing chronic pain in individuals (Grol-Prokopczyk 2017). Grol-Prokopczyk et al’s study identified large magnitude differences in chronic pain scores, with low income ethnic minorities experiencing greater pain compared to more wealthy non-minority individuals (Grol-Prokopczyk 2017).

Another critical factor to consider in chronic pain management is observations that both laypeople and medical professionals often hold the false belief that minorities have higher rates of pain which contributes to widely acknowledged racial disparities in the assessment and treatment of chronic pain (Trawalter and Hoffman 2015, Hoffman, Trawalter et al. 2016). However, reviews of the literature strongly suggest that racial and ethnic minorities are more sensitive to pain, with lower pain tolerance and higher pain ratings compared to non-Hispanic Whites (Kim, Yang et al. 2017). There is evidence that both hypervigilance (Campbell, Edwards et al. 2005) and adverse effects of racial discrimination (Edwards 2008, Goodin, Pham et al. 2013, Walker Taylor, Campbell et al. 2018) play important roles in higher reported pain in Black populations.

High prevalence of chronic pain, combined with adverse consequences of pain medication dependence, have increased interest in mindfulness-based interventions (Hilton, Hempel et al. 2017, Marske, Shah et al. 2020). Common interventions practiced for chronic pain reduction include mindfulness-based stress reduction (MBSR), mindfulness-based cognitive therapy (MBCT), and integrative treatments that combine mindfulness techniques with other conventional and complementary medicines. Mindfulness elements are postulated to allow individuals to reframe experiences such as chronic pain by refocusing the mind on the present and increasing awareness of external surroundings and inner sensations (Grant and Rainville 2009).

Mindfulness meditation is associated with statistically significant improvements in depression, physical and mental health, pain self-efficacy, and sleep (Morone, Greco et al. 2016, Turner, Anderson et al. 2016, Marske, Shah et al. 2020). However, the magnitude of health improvement can vary across studies. Cross-sectional cohort studies of people engaging in long-term meditation practices find that higher levels of mindfulness are associated with lower pain intensity ratings and changes in the structure of brain regions related to pain perception (Luders, Phillips et al. 2012). Yet, mindfulness intervention studies as a treatment for chronic pain have heterogeneous results overall. While some studies focus on specific types of pain such as low back pain (Cramer, Haller et al. 2012, Morone, Greco et al. 2016), fibromyalgia (Kozasa, Tanaka et al. 2012) or chronic migraines, others broadly target chronic pain. Systematic reviews (Hilton, Hempel et al. 2017, Majeed, Ali et al. 2018) indicate that treatment of chronic pain with mindfulness interventions has moderate success, and more comprehensive research is needed to support a recommendation for the use of mindfulness meditation for any chronic pain symptomology. In addition, despite the potential for effective pain management in minority populations which may suffer from elevated rates and levels of chronic pain, inclusion of minorities in mindfulness based interventions is relatively rare (Waldron, Hong et al. 2018).

Another increasingly common and effective non-pharmacological intervention for treating chronic pain is group medical visits. Within the environment of group medical visits, peer support from group members, an understanding of self-healing, and overall relationships are built with both the clinician and other participants (Parikh, Rajendran et al. 2019, Znidarsic, Kirksey et al. 2021). These experiences suggest that group medical visits can be a viable intervention for alleviating chronic pain. Our prior work evaluating racially and ethnically diverse patients with chronic pain during group medical visits over a 12-month course reported decreases in pain level, perceived stress, and depression as well as improvements in sleep quality (Gardiner, Dresner et al. 2014). Group medical visits have positive effects, with a 2017 study of a Mindfulness Based Stress Reduction program for 20 low-income minority adults with chronic pain and comorbid depression determining that group treatment contributed to enhanced coping with chronic pain and improvement in the sense of control over one’s health condition (Lestoquoy, Laird et al. 2017). Integrative medical group visits (IMGV), which focus on increasing access to integrative health care, have also been particularly effective in managing and treating chronic conditions (Thompson-Lastad, Gardiner et al. 2019, Znidarsic, Kirksey et al. 2021). They can be used as adjuncts to medications, particularly in underserved diverse, low-income populations with limited access to non-pharmacological therapies. There are benefits to both the patients and clinicians participating in integrative group medical visits, with improvements in patient physical and mental health (Thompson-Lastad, Gardiner et al. 2019).

Our recent randomized control trial (RCT) assessed the effectiveness of IMGV compared to a Primary Care Provider (PCP) visit in patients with chronic pain and depression in a 9-week single-blind two arm randomized control trial with a 12-week maintenance phase (intervention-medical groups; control-primary care provider visit) (Gardiner, Luo et al. 2019). All participants (N=155) were randomized (1:1) to either intervention (IMGV) or control group. There were no group differences in average pain level in predominantly low income racially diverse adults with nonspecific chronic pain and depressive symptoms. However, the IMGV group had fewer emergency department visits at 9 weeks, and this group reported reduced pain medication use at 21 weeks compared to controls. Further analyses identified and characterized a predictive relationship between depression and chronic pain interference. This prediction was mediated by high perceived stress, low pain self-efficacy, and poor sleep quality, potential targets for attenuating the adverse effects of depression on functional outcomes (Nephew, Incollingo Rodriguez et al. 2021).

The objective of the present study was to assess the effects of race on the primary pain outcomes of the IMGV RCT. It was hypothesized that there would be racial differences in the effects of IMGV on pain related outcomes. Related findings could enhance our understanding of treatment outcomes in diverse patient populations participating in similar complementary interventions.

## METHODS

### Study Setting

The study was approved by the Boston University Medical Campus Institutional Review Board (IRB) and the community health center’s (CHC) research committees (IRB Approval Number: H33096). We registered this randomized controlled trial (RCT) in the international trial register [ClinicalTrials.gov: Identifier NCT02262377]. The RCT was conducted at an ambulatory primary care clinic in the outpatient building at Boston Medical Center (BMC) in Boston MA and at two community health centers: Codman Square Health Center (CSHC) and Dorchester House Health Center (DHHC). BMC and the two affiliated federally qualified health centers serve income-disadvantaged, racially diverse and ethnically diverse populations with health disparities in the treatment of chronic pain and depression. BMC is a private, not-for-profit, academic medical center. DHHC is a 3-story modern health facility with 40 clinical exam rooms. DHHC has approximately 30,000 active patients. CSHC has 22 exam rooms, and three new rooms for group medical visits. CSHC has approximately 21,000 active patients. These locations were chosen because they are the setting where there is a high prevalence of income-disadvantaged diverse patients with chronic pain and depression who do not have access to non-pharmacological treatments. All three sites were accessible by public transportation.

Participants were recruited through their clinicians’ outpatient referral, clinicians’ letter to patients about the study, or self-referral. After being contacted by the research assistant (RA), patients then consented to be screened. Our inclusion criteria included: age 18 years or older, able to communicate in English language, score of ≥ 5 on the Patient Health Questionnaire-9 (PHQ-9), score of ≥ 4 on a 0–10 scale measuring daily chronic pain intensity for at least 12 weeks, and having a PCP located at the site where the IMGV was being held. The exclusion criteria included: self-reported symptoms of psychosis or mania, active substance abuse (alcohol, cocaine or heroin use in the last 3 months), previous participation in an IMGV, a new pain treatment in the past month or plans to begin any new pain treatments in the next three months, active suicidality, any other severe disabling chronic medical or psychiatric co-morbidities preventing attendance to the IMGV, or no access to the internet during the study period. If the eligibility was verified and there was patient written consent, the patient was enrolled in the study.

### IMGV intervention

The IMGV intervention included three concurrent deliveries of the same self-management curriculum delivered with different formats–an in-person MGV, and two adjunct companion technologies available on a computer tablet provided to the intervention participant. The first technology was the Our Whole Lives (OWL), an e-Health toolkit platform, and the second technology was an Embodied Conversational Agent (ECA). A detailed description of the IMGV self-management intervention has previously been described [34]. The IMGV consisted of a total of ten in-person medical group visits each lasting 2.5-hours conducted weekly from week 1 to week 9 (9 in-person sessions plus OWL/ECA). This is followed by a 12-week maintenance phase where there is access to the technology only (OWL/ECA). A tenth and final in-person session is conducted at week 21.

At the beginning of each session of the IMGV, participants measured their vital signs, mood, and pain levels. They then met individually with a trained physician (a co-facilitator) for a medical assessment. A Black certified MBSR facilitator (see below) then led mindfulness practices adapted from Mindfulness Based Stress Reduction (meditation, body scan, etc.). Patients were instructed in the principles of mindfulness and other self-management techniques (such as acupressure and self-massage). Each week, the physician facilitated a discussion on health topics such as stress, insomnia, depression, chronic pain cycle, activity, and healthy food choices. Finally, the IMGV ended with a healthy meal, which mirrored the healthy nutrition topic in each session. In addition to a physician, an experienced co-facilitator with training in mindfulness (certified MBSR instructor, yoga, and meditation teacher) attended all groups. Facilitators were mentored via direct observation of two pilot group visits, one-on-one meetings, and phone calls by an experienced MBSR trained faculty.

To reinforce all content delivered in the in-person group, an internet-based platform called Our Whole Lives; an electronic health toolkit (OWL) delivered the same in-person curriculum. OWL could be accessed with a computer, smart phone, or tablet. The ECA, an automated female character, emulated the conversational behavior of an empathic coach [48]. The ECA (Gabby) reviewed all the content discussed in the IMGV with the participants outside of the in-person group. A Dell Venue 8 Pro tablet was distributed to all intervention participants in the first session of the group.

After the nine-week in-person group visit phase concluded, the intervention participants entered a 12-week maintenance phase. The intervention participants retained the study tablet and continued to have access to the ECA and the OWL website. At the end of the 21 weeks, there was one final in-person group visit. A trained study RA directly observed all groups and assessed the facilitator’s adherence to the intervention components through a monitoring and evaluation checklist. These checklists were used to assess each IMGV session at all sites during the study.

### Control Condition

All participants randomized to the control group were asked to visit their PCP during the study period (baseline to 21 weeks). We verified a PCP clinical visit via electronic medical record (EMR) documentation. We did not collect data on the duration or content of the visit.

### Clinical Outcomes

Outcome data were collected at baseline, 9 weeks, and 21 weeks. Baseline demographic data included: age, gender, race, ethnicity, income, work status, and education. Our primary outcomes included in the present analysis consist of: 1) self-reported pain measured by the Brief Pain Inventory [(BPI) pain interference, pain severity [27] and 2) depression level measured by the PHQ-9, a self-reported depression screen [28]. BPI pain interference, and pain severity are on a 0-10-point scale. The higher the score, the more severe the pain. PHQ-9 is on a 0-27-point scale. The higher the score, the more severe the depression.

Secondary outcomes included were perceived stress. The Perceived Stress Scale (PSS) is a self-reported questionnaire designed to assess “the degree to which individuals appraise situations in their lives as stressful [29].”

### Analytic Plan

One-way ANOVAs were performed to evaluate differences in separate pain outcome values by participant race at each time point. Pain outcome values (interference, severity) were then each analyzed separately using a 2(condition)x2(race)x3(time) mixed analysis of variance, with repeated measures at baseline, nine weeks, and 21 weeks on the third factor. That is, the interaction between study condition and participant race was tested in relation to the trajectory of pain outcome values across the study observation period.

In these analyses, variables between only Black (58%) and White (19%) participants were accessed, which excludes 36 (23%) participants who indicated “other.” Due to the heterogenic nature of the “other” including 21 Hispanic, 10 mixed-race, 2 Asian, 2 Caribbean, and 1 Arab, we chose to dichotomize the analysis. Further information regarding these participants is available in previous publications (Gardiner, Luo et al. 2019, Nephew, Incollingo Rodriguez et al. 2021). T-tests were also conducted between race variables (Black and White) and demographic information (Table 1).

**Table 1.** Demographics split by race variables.

|  | Black (N=90) |  | White (N=29) |  | T-test | p-value |
| --- | --- | --- | --- | --- | --- | --- |
| Age (22-84) | $\mu=50.5(sd=1.3)$ | | $\mu=55.9(sd=2.1)$ | | 2.12 | **0.018\*** |
| Gender | **n** | **%** | **n** | **%** | 0.82 | 0.21 |
| *Female* | 75 | 83 | 26 | 90 |  |  |
| *Male* | 15 | 17 | 3 | 10 |  |  |
| Income | **n** | **%** | **n** | **%** | 0.13 | 0.45 |
| *Less than \$5K* | 16 | 18 | 2 | 7 | | |
| *\$5K-\$29.99K* | 41 | 45 | 18 | 62 | | |
| *\$30K and over* | 7 | 8 | 3 | 10 | | |
| *Refused/DK/No personal income* | 26 | 29 | 6 | 21 |  |  |
| Work Status | **n** | **%** | **n** | **%** | -2.23 | **0.014\*** |
| *Full/Part time* | 19 | 21 | 4 | 14 |  |  |
| *Unemployed* | 14 | 16 | 1 | 3 |  |  |
| *Retired/Homemaker* | 11 | 12 | 3 | 10 |  |  |
| *Sick Leave/Disability* | 40 | 44 | 15 | 52 |  |  |
| *Other* | 6 | 7 | 6 | 21 |  |  |
| Education Level | **n** | **%** | **n** | **%** | -1.52 | **0.06** |
| *Less than high school/Some HS* | 15 | 17 | 5 | 17 |  |  |
| *High school degree* | 37 | 41 | 6 | 21 |  |  |
| *Some college/AA degree* | 31 | 34 | 13 | 45 |  |  |
| *College degree or higher* | 7 | 8 | 5 | 17 |  |  |
| Comorbidity Profile Among Participants** | $\mu=2.6(sd=0.2)$ | | $\mu=1.8(sd=0.3)$ | | 2.48 | **0.007\*** |
|  | **n** | **%** | **n** | **%** |  |  |
| *No comorbidities* | 13 | 14 | 8 | 28 |  |  |
| *1 comorbidity* | 13 | 14 | 7 | 24 |  |  |
| *2 comorbidities* | 12 | 13 | 6 | 21 |  |  |
| *3 comorbidities* | 27 | 30 | 3 | 10 |  |  |
| *4 comorbidities* | 11 | 12 | 1 | 3 |  |  |
| *5 or more comorbidities* | 14 | 16 | 4 | 14 |  |  |
| Attendance of IMGV Participants<br>(Number of intervention sessions<br>attended) (0-10) | n = 45 (IMGV<br>patients)<br>$\mu=6.2$ (sd=0.5) | | n = 13 (IMGV<br>patients)<br>$\mu=5.5$ (sd=0.8) | | 0.77 | 0.22 |
| Patient Health Questionnaire (PHQ-9)<br>(0-27) |  |  |  |  |  |  |
| *Baseline* | $\mu=11.9$ (sd=0.6) | | $\mu=11.8$ (sd=1.1) | | -0.11 | 0.46 |
| *9 weeks* | $\mu=10.0$ (sd=0.6) | | $\mu=10.6$ (sd=1.1) | | 0.43 | 0.33 |
| *21 weeks* | $\mu=9.8$ (sd=0.6) | | $\mu=9.9$ (sd=1.3) | | 0.12 | 0.45 |
| Perceived Stress Scale (PSS-4) (0-16) |  |  |  |  |  |  |
| *Baseline* | $\mu=7.8$ (sd=0.4) | | $\mu=7.0$ (sd=0.6) | | 1.11 | 0.13 |
| *9 weeks* | $\mu=6.7$ (sd=0.4) | | $\mu=7.0$ (sd=0.7) | | -0.36 | 0.36 |
| *21 weeks* | $\mu=6.4$ (sd=0.4) | | $\mu=6.8$ (sd=0.6) | | -0.44 | 0.33 |

Note. $\mu$ = mean; sd = standard deviation
\*denotes significance ( $p<0.05$ )
\*\*Comorbidity Profile includes hypertension, obesity, insomnia, diabetes, vitamin D deficiency, PTSD, and substance use disorders (alcohol, tobacco, opioid, cocaine, marijuana)

## RESULTS

Three hundred forty-three patients were assessed for eligibility, 209 patients were eligible, and 159 were enrolled and randomized to intervention (80) or control. The consort diagram and baseline characteristics were previously published (Gardiner, Luo et al. 2019).

Of the 155 participants who all have baseline chronic pain and depression, the average age was 51 years old, 86% identified as female, 58% identified as Black, 23% identified as “other” race, and 19% identified as White. Sixty three percent earned less than $30,000 a year, only 21% worked full or part time, 14% were unemployed and 42% were on work disability. Common co-morbidities in the participants were hypertension (41%), insomnia (26%), anxiety (28%), Post-Traumatic Stress Disorder (PTSD) (16%), and any substance use disorder (25%). The average PHQ-9 score for depressive symptoms was ∼ 12 at baseline, which is characterized as moderate depression.

T-tests between race (Black and White) were run based on categorical scoring of various demographics variables. Significant differences were observed in age (p=0.018), work status (p=0.014), and comorbidity profile (p=0.007), and a trend was observed in education (p=0.06). The average ages of race variables were μ=50.5 for Black patients and μ=55.9 for White patients. These averages are within the same age group and were not a factor in subsequent analyses. Work status between race suggested that more Black patients (21%) were full time employed compared to White patients (14%). Black patients reported unemployment at 16% and White patients reported unemployment at 3%. However, given the group sizes based on race and the large percentage of White patients (21%) citing ‘other’ as work status, it is difficult to evaluate the role of works status in this study.

Across treatment conditions, at baseline, Black participants showed no difference in pain interference at baseline, but they did show significantly higher pain severity than White participants. This same pattern was observed at nine and 21 weeks. Additionally, at nine weeks, Black participants also showed marginally higher pain interference than White participants. See Table 2.

**Table 2.** Descriptive statistics and ANOVA results of pain differences by race.

|  | White Participants |  | Black Participants |  | *F* | *df* | *p*-value |
| --- | --- | --- | --- | --- | --- | --- | --- |
|  | Mean | *SD* | Mean | *SD* |  |  |  |
| Pain interference |  |  |  |  |  |  |  |
| Baseline | 5.93 | 2.18 | 6.35 | 2.30 | 0.52 | 1, 123 | .471 |
| 9 weeks | 4.61 | 2.49 | 5.80 | 2.56 | 3.66 | 1, 110 | **.058** |
| 21 weeks | 4.78 | 2.57 | 5.31 | 2.71 | 0.82 | 1, 112 | .366 |
| Pain severity |  |  |  |  |  |  |  |
| Baseline | 5.60 | 1.84 | 6.69 | 1.85 | 6.88 | 1, 123 | **.010** |
| 9 weeks | 5.10 | 1.82 | 6.03 | 2.16 | 4.38 | 1, 110 | **.039** |
| 21 weeks | 4.84 | 1.63 | 5.83 | 2.21 | 4.01 | 1, 112 | **.048** |

*df* = degrees of freedom

A 2×2×3 mixed analysis of variance revelaed that while pain interference did change over time, this trajectory was not related to the interaction between treatement condition and race. See Table 3. However, pain severity differed significantly over time and there was also a significant three-way interaction among severity, treatment condition, and race. This test did not violate the assumption of sphericity, *p* = .529. See Table 3 for test statistics per interaction and Figure 1 for trajectories over time.

**Table 3.** Pain interference and severity over time by treatment condition and race.

|  | *F* | *df* | *p*-value |
| --- | --- | --- | --- |
| Interference (baseline, 9 weeks, 21 weeks) | 10.69 | 2 | **<.001** |
| Interference x race | 1.73 | 2 | .179 |
| Interference x treatment | 1.11 | 2 | .329 |
| Interference x race x treatment | 0.11 | 2 | .885 |
| Severity (baseline, 9 weeks, 21 weeks) | 10.40 | 2 | **<.001** |
| Severity x race | 0.28 | 2 | .759 |
| Severity x treatment | 1.17 | 2 | .438 |
| Severity x race x treatment | 3.28 | 2 | **.039** |

*df* = degrees of freedom

**Figure 1.**
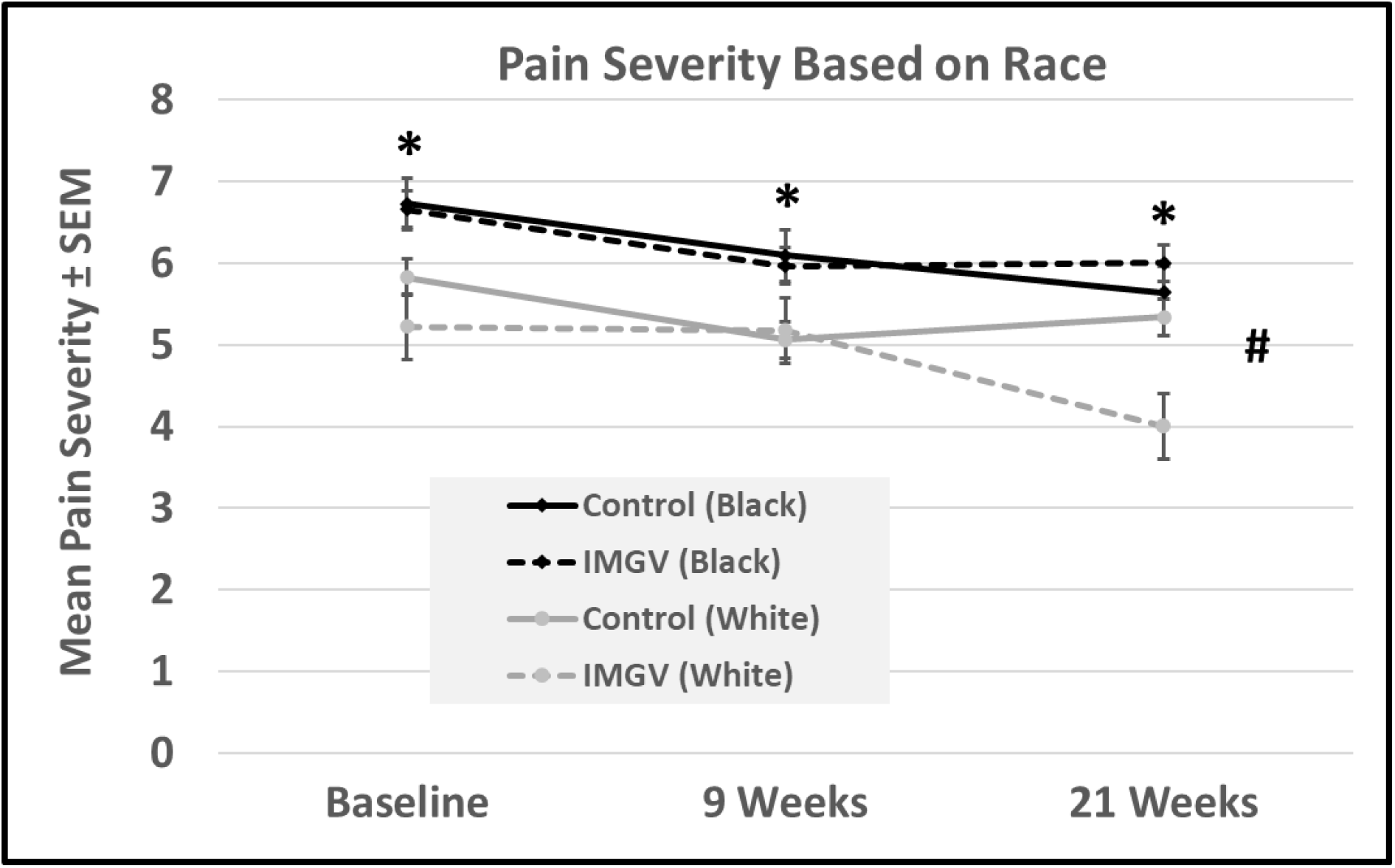
Pain Severity Across Treatment Groups. ^*^ denotes differences by race at each timepoint (p<0.05). # denotes significant overall interaction between pain severity, treatment condition and race (p<0.05)

## DISCUSSION

The current analysis identified significant and substantial group-based differences in the effects of a mindfulness intervention for chronic pain. Black subjects experienced higher pain severity over the 21 week study period compared to White subjects. Where IMGV reduced pain severity in White subjects, it had no effect in Black subjects. While many complementary interventions for chronic pain are generally effective, there is a critical need for a greater understanding of their efficacy in minority populations.

Significant differences for comorbidity profile between Black and White patients were identified in the current study, with Black patients more likely to suffer from multiple diagnoses. Three or more comorbidities were observed in 58% of Black participants compared to 28% of White participants. Racial differences in comorbidity profiles have been associated with similar findings in recent studies, with Black patients suffering higher proportions of comorbidities compared to White patients (Kabarriti, Brodin et al. 2020). These racial variations in comorbidities may also contribute to decreased utilization of treatment (Hankerson, Fenton et al. 2011). While education level did not differ between Black and White patients, it is worth noting the trend that Black patients participated less in ‘some college/AA degree’ and ‘college degree or higher’ compared to that of White patients. This trend could be related to digital literacy competencies in healthcare, where Black patients are disproportionally affected by these challenges (Smith and Magnani 2019).

Despite the increased need for effective pain management and an intervention targeted at high-risk Black communities, there are substantial challenges to complementary interventions for these groups. The initial analysis by race at each timepoint revealed that Black subjects had higher pain severity at baseline and 9 and 21 weeks later, similar to findings from other interventions (Meints, Wang et al. 2018). This increased severity may be due to other factors including stress levels (Sternthal, Slopen et al. 2011), poor mindfulness class attendance (Palmer, Rivers et al. 2021), or lack of mindfulness practice at home (Webb, Khubchandani et al. 2019). Cultural minority experiences and perceptions could also contribute to the lack of IMGV effect on reducing pain severity in these patients (Johnson, Saha et al. 2004, Shepherd, Willis-Esqueda et al. 2018). It is noted that we have not accounted for income, stress level, or housing insecurity, which may also play roles in how minority populations respond differently to pain as well as treatment. However, several differences in comorbidities and demographics were observed in the present population.

The present report of racial differences in pain support the consistent documentation of racial disparities (Janevic, McLaughlin et al. 2017), where Black patients often suffer from the greatest levels of chronic pain and pain related adverse outcomes (Meints, Cortes et al. 2019). These disparities extend to pain management as well, where false beliefs and implicit bias in healthcare may drive racial disparities in the assessment and treatment of pain (Hoffman, Trawalter et al. 2016, Meints, Cortes et al. 2019, Morales and Yong 2021). Racial disparities in chronic low back pain are associated with greater pain sensitivity in Black patients compared to non-Hispanic Whites (Meints, Wang et al. 2018), and this enhanced sensitivity was partially mediated by pain catastrophizing. Increased focus on racially relevant aspects and catastrophizing in similar interventions may attenuate racial disparities in pain management.

There are two key elements, one in the control groups, one in the treatment groups, to the differential trajectories in pain severity underlying the interaction between pain severity, treatment, and race. Both Black and White control subjects reported decreased severity over the first 9 weeks, which could be related to the required GP visit and/or an effect of enrolling in the study. While this decreasing trend continued in the Black control subjects, pain severity plateaued in White control subjects. In the IMGV treatment groups, severity plateaued in Blacks between 9 and 21 weeks during the maintenance phase, where it decreased in White subjects. The combination of these differences in trajectories, combined with the overall higher pain severity in Black, contributed to the interaction between severity, treatment, and race. A potential explanation for the pattern in controls is that that control condition of seeing a GP during the course of the study had a more sustained effect on decreasing pain severity in Black subjects. Although it is unclear why, the data from the IMGV groups suggests increased efficacy in White subjects, specifically during maintenance phase from 9 to 21 weeks. Given the significant effect of IMGV on reducing pain medication use in the overall study population, this factor was investigated, but there were no race-based differences or interactions between medication use, treatment, and race.

A meta-analysis of the effects of race in mindfulness and acceptance based interventions for depression reported no moderation by race (Dawson, Jones et al. 2022), contrasting with the present study. This is particularly noteworthy given the prior report of the predictive relationship between depression and chronic pain in the IMGV data (Nephew, Incollingo Rodriguez et al. 2021). This could indicate that aspects of the group medical visits and/or the maintenance phase protocols were key factors in the race based difference in pain severity trajectories. The presence of the greater divergence in trajectories during the maintenance phase, which involved interaction with the ECA and OWL website (and related digital health literacy), suggests that differential responding during this period was the more substantial factor.

There is growing concern about digital health literacy accentuating current racial disparities in healthcare (Smith and Magnani 2019). Black and Latino populations are disproportionally affected by challenges in health literacy in general, and racial minorities are less likely to use patient portals, even when adjusting for educational attainment (Gordon and Hornbrook 2016). A 2022 systematic review of qualitative studies of enablers and barriers to telehealth interventions specifically for people with chronic pain concluded that health literacy was a critical barrier (Fernandes, Devan et al. 2022). Encouragement of self-efficacy and empowerment of patients were associated with successful telehealth interventions for chronic pain management. It is possible that racial differences in the usage and/or efficacy of the ECA and OWL website led to the racial differences in overall treatment efficacy.

Another hypothesis for the racial difference in the response to IMGV is that this intervention is more effective at treating lower pain severity, where higher severity requires other treatments, such as opiates. Complementary, non-pharmacological treatments tend to be more effective for mild and moderate pain compared to higher levels, which are more likely to respond to opiate based interventions. The greater severity also indicates levels that may be more resistant to treatment in general (Borsook, Youssef et al. 2018).

The present results, both the overall differences by race at each timepoint and the divergent trajectories, may also have been mediated by pain catastrophizing. Greater catastrophizing by Blacks is associated with higher pain severity (Meints, Miller et al. 2016, Dorado, Schreiber et al. 2018) and the lack of pain attenuation in Black subjects in the present study may be related to relatively greater catastrophizing during the maintenance phase. The effects of mindfulness interventions are complex and mixed, where some specific aspects (Observe) reduce the adverse impact of catastrophizing on pain and others (Non-judgement and Awareness) accentuate these impacts. Careful consideration and implementation of particular facets of mindfulness in customized pain interventions, rather than the use of standardized mindfulness protocols, may improve outcomes for all groups. There is clearly potential for interactions between several factors, including digital health literacy and pain catastrophizing, to affect the levels and trajectory of pain severity in the present and similar studies.

### Limitations

There are several limitations of the present analysis, such as the lack of inclusion of other races due to insufficient sample sizes and the potential for obscuring other important differences due to stratifying by race. We were also unable to assess racism or experiences of discrimination. A final limitation is the lack of data on participant digital health literacy levels, where racial differences in this factor may have affected the results. Suggestions for future research include larger, more diverse study samples, more comprehensive assessment of catastrophizing and digital health literacy, and the inclusion of longitudinal manipulative studies of omitted factors to determine causality.

## CONCLUSION

The present analysis indicates that overall efficacy in the associated RCT of IMGV differed by race. Black subjects suffered from higher pain severity throughout the 21 weeks of the study, yet were less likely to beneficially respond to the pain intervention compared to White subjects. Potential explanations for this disparity which require further investigation include differential comorbidity profiles, catastrophizing, and digital health literacy. Racial disparities in pain affect not only the experience of pain, but also how individuals respond to complimentary interventions. Due to the urgent need for effective pain management interventions targeted at high risk minority populations, increased emphasis should be directed towards racially relevant aspects of both pain and the response to complimentary interventions to address disparities.

## Data Availability

Correspondence concerning this article should be directed to Paula Gardiner.

